# Impact of COVID-19 on Mental Health: A Longitudinal Study Using Penalized Logistic Regression

**DOI:** 10.1101/2021.02.21.21252159

**Authors:** Jingyu Cui, Jingwei Lu, Yijia Weng, Grace Y. Yi, Wenqing He

## Abstract

The COVID-19 pandemic has posed significant influence on the public mental health in a stealthy manner. Current efforts focus on alleviating the impacts of the disease on public health and economy, with the psychological effects due to COVID-19 largely ignored. In this paper, we analyze a mental health related dataset from the US to enhance our understanding of human reactions to the pandemic. We are particularly interested in providing *quantitative* characterization of the pandemic impact on the public mental health, on top of *qualitative* explorations. We employ the multiple imputation by chained equations (MICE) method to deal with missing values and take the logistic regression with least absolute shrinkage and selection operator (Lasso) method to identify risk factors for mental health. The analyses are conducted to a large scale of online survey data from 12 consecutive weeks, so that the longitudinal trend of the risk factors can be investigated. Our analysis results unveil evidence-based findings to identify the groups who are psychologically vulnerable to COVID-19. This study is useful to assist healthcare providers and policy makers to take steps for mitigating the pandemic effects on public mental health.

## 1 Introduction

Since the outbreak of the COVID-19 pandemic, people’s life style has been changed significantly. Isolation and social distancing have been broadly implemented, and virtual social interactions have been encouraged. These changes have presented great challenges to people’s work, study and living. Furthermore, they considerably affect people’s psychological reactions to the disease, with more occurrence of emotional distress and social disorder during, and probably after, the outbreak. Despite these facts, no sufficient resources have been available to manage or attenuate the pandemic effects on mental health and well-being (Taylor, 2019). Psychological reactions to the pandemic typically include maladaptive behaviours, emotional distress, and defensive responses (Taylor, 2019). Individuals with psychological issues are especially vulnerable, and those who are unable to adjust to the new life style become prone to mental health issues.

A number of studies have been conducted to investigate how the COVID-19 pandemic may affect people psychologically. Cao et al. (2020) conducted a survey on college students in China and showed that more than 24% of the students were experiencing anxiety. Moreover, living in urban areas, family income stability and living with parents are protective factors against anxiety, and having relatives or acquaintances infected with COVID-19 increases anxiety. Spoorthy et al. (2020) investigated the mental health problems faced by healthcare workers during the COVID-19 pandemic. Kang et al. (2020) conducted a cross-sectional study on 994 medical and nursing staffs in Wuhan, China. They found that 36.9% of the study subjects had subthreshold mental health disturbances, 34.4% had mild disturbances, 22.4% had moderate disturbances, and 6.2% had severe disturbances. Cai et al. (2020) carried out a cross-sectional observational study including doctors, nurses, hospital staffs throughout Hunan province of China between January and March 2020; they reported that the medical staffs experienced emotional stress during the COVID-19 pandemic.

While those studies provided descriptive results by summarizing the information obtained from the questionnaire which are typically collected at a certain time point, important questions remain unanswered. Notably, it is unclear how the impact of COVID-19 changes over time; what factors are relevant to describe the impact of the pandemic; and how the severity of the pandemic is quantitatively associated with the risk factors. In this paper, we examine these questions and aim to provide quantitative insights. Our explorations are carried out using a large scale online public survey database from *U*.*S. Census Bureau*. The data include twelve datasets with different sizes collected over 12 consecutive weeks from April 23, 2020 to July 21, 2020, in which the smallest dataset contains 41,996 subjects and the largest dataset has 132,961 individuals. The participants in the survey age from 18 to 88 and come from the 50 states and Washington, D.C. in the US. The survey includes multiple questions perceived to be relevant to understand the impact of the pandemic on the public. To quantitatively identify the risk factors for impacting the mental status by the pandemic, we engage penalized logistic regression, specifically with the least absolute shrinkage and selection operator (Lasso) method (Tibshirani, 1996), to conduct simultaneous variable selection and parameter estimation. However, a direct application of the Lasso method is not possible for the data because they have missing observations with the rates ranging from 12.8% to 14.5% over the 12 weeks. To overcome this issue, we employ the multiple imputation by chained equations (MICE) (Raghunathan et al., 2001; Yu et al., 2007) to impute missing values. Further, survey data commonly involve measurement error due to recall bias, inability of providing precise descriptions of some answers, and reporting errors, it is imperative to address this issue when pre-processing the data. To this end, we combine the levels of those highly related categorical variables to mitigate the measurement error effects.

The remainder of the manuscript is organized as follows. Section 2 introduces the data and describes how the data is pre-processed. Section 3 discusses the MICE method and reports the features of the resultant imputed data. In Section 4, the Lasso framework under logistic regression is introduced. In Section 5, we apply the Lasso logistic regression to analyze the pre-processed data and report the findings. Lastly, we conclude the paper with discussion in Section 6.

## 2 Mental Health Data

### 2.1 Raw Data

The data used in this project are from phase 1 of the Household Pulse Survey, conducted within 12 weeks from April 23, 2020 to July 21, 2020 by the U.S. Census Bureau (https://www.census.gov/). The survey aims to study the pandemic impacts on the households across the US from social and economic perspectives. The participants of the survey come from the 50 states and Washington, D.C. of the US, aging from 18 to 88. The gender ratio (the ratio of males to females) remains fairly stable ranging between 0.6 and 0.7 over the 12 weeks. Figure 1 displays the counts of the participants by the status of their states which are classified into four categories according to the severity of the pandemic: *mild, moderate, large daily increase*, and *serious*; such a classification is conducted based on the trend of the cumulative cases over time, as shown in Figure 2 which is generated by the data from the *Centers for Disease Control and Prevention* (https://data.cdc.gov/). The states whose curves of the number of cumulative cases are on the very top are classified in the class of *serious*. The states whose curves of the number of cumulative cases stay at the bottom are regarded as the mild pandemic states. In-between, the states with steep curves are taken to be in the class with large daily increases, and the states with less steep curves are regarded as the states with moderate daily increases. It is seen that the majority (72.5%) of the participants come from the states with mild pandemic and the least proportion (2.3%) of subjects are from the states with a serious pandemic. Table 1 lists the state members for each category.

**Table 1:**
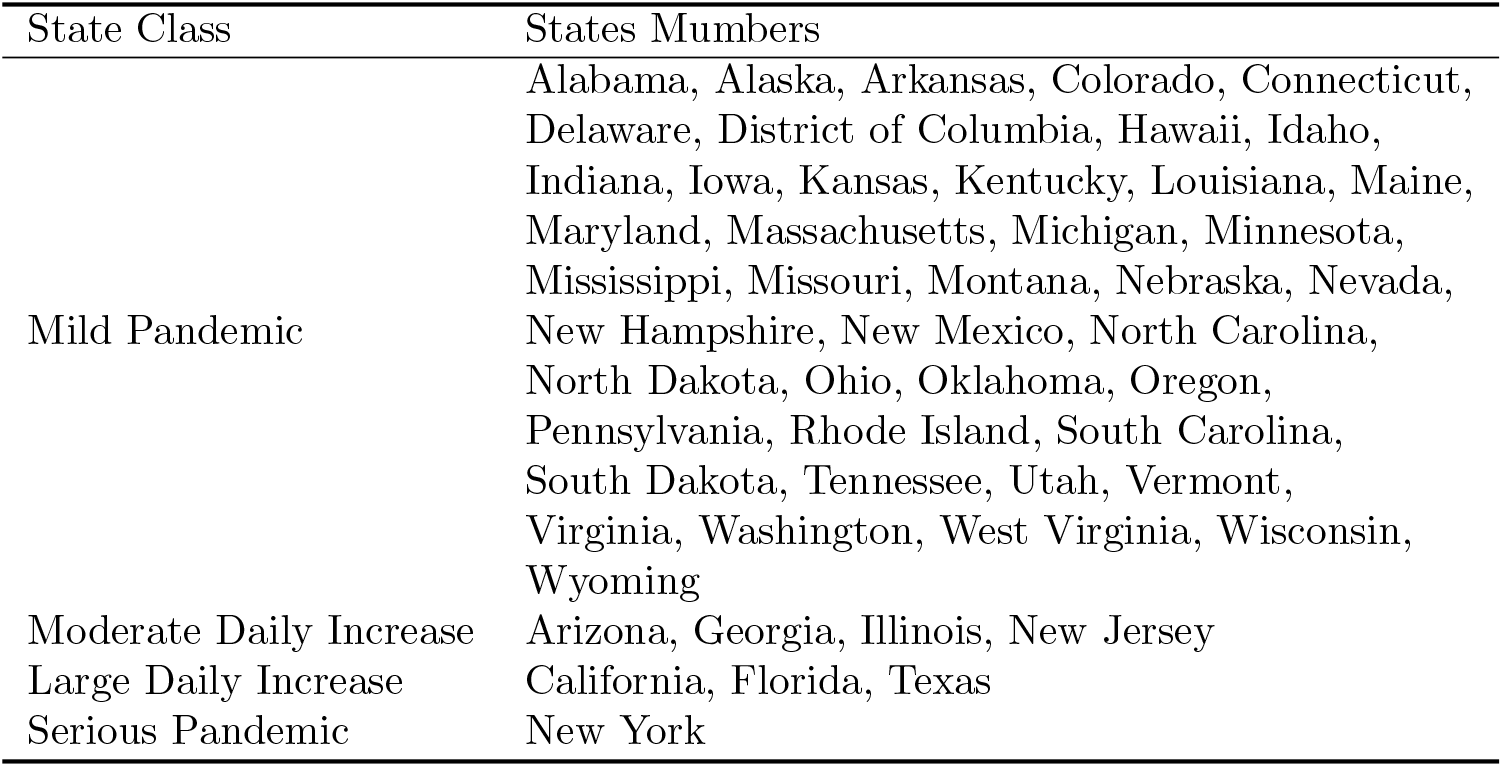
Members of four state classes

**Figure 1:**
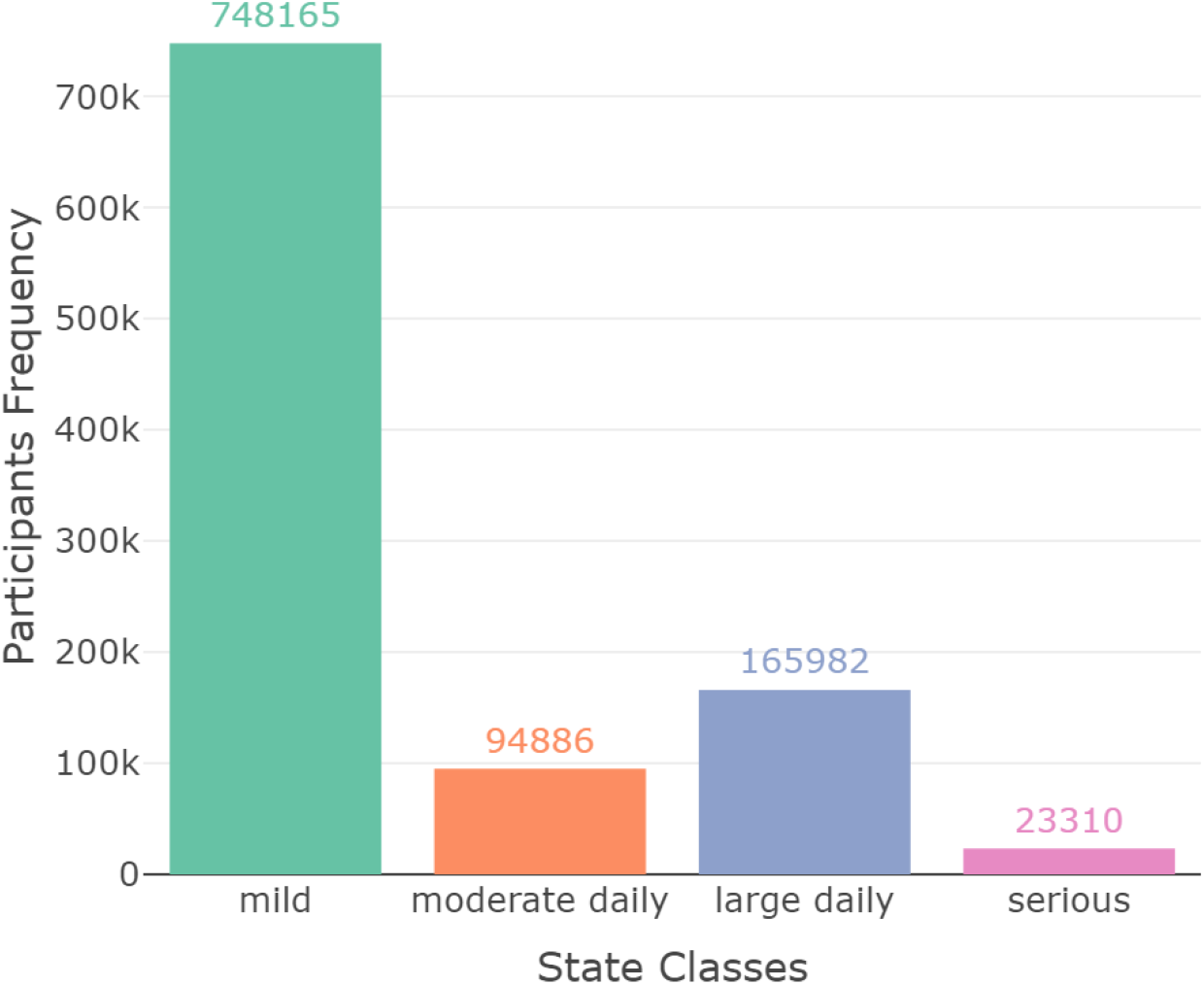
The number of the participants in the four classes classiffied by the severity of the pandemic

**Figure 2:**
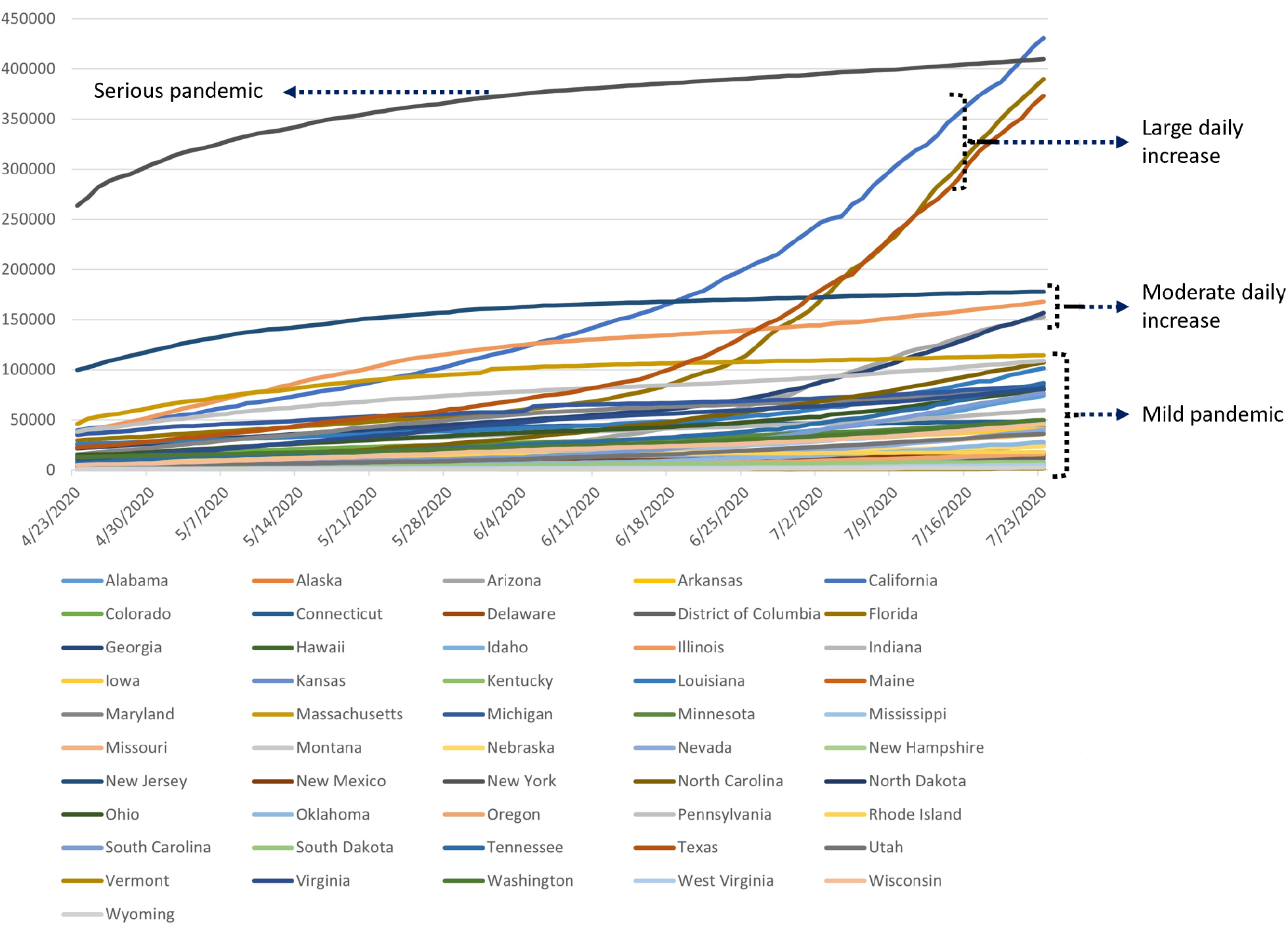
The number of cumulative cases for the 50 states and Washington, D.C. of the US over time

The survey contains 50 questions ranging from education, employment, food sufficiency, health, housing, social security benefits, household spending, stimulus payments, to transportation. Nine questions, such as *“Where did you get free groceries or free meals”* and *“How often is the Internet available to children for educational purposes”*, are not included in the analysis because they are not perceived as sustainable factors affecting mental health. Some of the questions are considered highly related to others. Specific manipulations have been done to remove possible correlations among those questions and to further reduce the measurement error, as discussed in the next subsection.

### 2.2 Pre-processing the Data to Reduce Errors

The survey is conducted on a weekly basis for 12 consecutive weeks, giving rise to 12 datasets each for a week. Missing observations and measurement error are typical features involved in the datasets. Before we conduct a formal analysis of the data, we implement a pre-processing procedure to mitigate the effects due to missingness and measurement error. In Section 3, we describe the steps of handling missing observations. Here we pre-process error-prone data to reduce the measurement error effects by combining questions to create new variables or by collapsing levels of variables to form binary variables.

Four questions in the survey measure people’s mental health status concerning four aspects: anxiety, worry, loss of interest, and feeling down. Each of them is a four-level Likert item (Joshi et al., 2015) measuring the frequency (1: Not at all; 2: Several days; 3: More than half the days; 4: Nearly every day) of each aspect happening during the past 7 days prior to the survey time. Using the middle point of those values as the threshold, we combine the four variables as a single binary response to reflect the mental health status of an individual. An individual is regarded to have mental health issues if the average of the four variables is greater than 2.5, and not otherwise.

Two variables describe the loss of work: *Wrkloss* indicates whether an individual in the household experiences a loss of employment income since March 13, 2020; *Expctloss* indicates if the individual expects a member in the household to experience a loss of employment income in the next 4 weeks because of the COVID-19 pandemic. These two variables are combined to form a single indicator which is denoted *Wrkloss*, with value 1 indicating that at least one of these two events happens. Two ordinal variables, *Prifoodsuf* and *Curfoodsuf*, are used to describe the food sufficiency status before the pandemic and at present, respectively. The *Foodcon*.*change* variable is constructed by comparing the current and the previous food sufficiency status, which is a binary variable taking 1 if the current food sufficiency status is no worse than the food status before the pandemic, and 0 otherwise. Variable *Med*.*delay*.*notget* is combined from two indicator variables *Delay* (indicating if medical care is delayed) and *Notget* (indicating if the medical care is not received), taking value 1 if either medical care is delayed or no medical care is received, and 0 otherwise. Predictor *Mort*.*prob* is combined from one binary variable and an ordinal variable, taking 1 if a participant does not pay last month’s rent or mortgage or does not have enough confidence in paying the next rent or mortgage on time, and 0 otherwise. In addition, three ordinal variables, *Emppay, Healins* and *Schoolenroll*, are modified by collapsing their levels to form binary categories. *Emppay* has value 1 if he/she gets paid for the time he/she is not working, and 0 otherwise. *Healins* has value 1 if an individual is currently covered by the health insurance, and 0 otherwise. *Schoolenroll* has value 1 if there is a child in the household enrolled in school, and 0 otherwise. Except for the variables discussed above, the remaining variables are kept as the original.

The final data include the binary response (indicating the mental health status of an individual) and 25 predictors measuring various aspects of individuals. To be specific, nine predictors show basic information: *State, Age, Male, Rhispanic, Race, Educ, Maritalstatus, Numper* (the number of people in the household), and *Numkid* (the number of people under 18 in the household); five varaibels concern the income and employment: *Income, Wrkloss, Anywork, Kindwork*, and *Emppay*; five variables are related to food: *Foodcon*.*change, Freefood, Tspndf ood, T spndprpd*, and *Foodcon f* ; three variables pertain to health and insurance: *Hlthstatus, Healins*, and *Med*.*delay*.*notget*; one variable, *Mort*.*prob*, is for mortgage and housing; and two variables, *Schoolenroll* and *Ttch Hrs*, reflect child education. The variable dictionary for the pre-processed data is shown in Table 2.

**Table 2:**
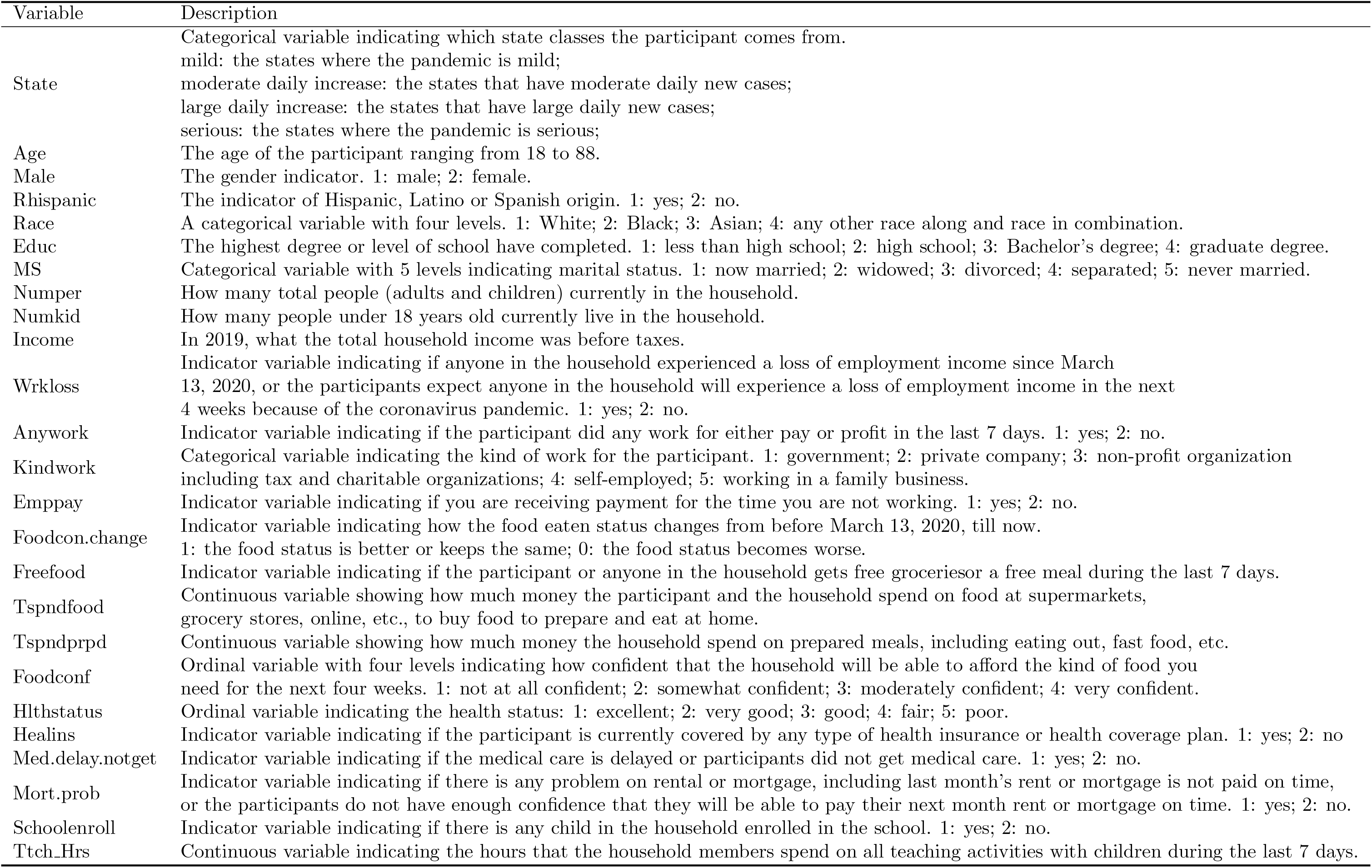
Variable Description

## 3 Accommodation of Missing Observations

The missing rates for all the pre-processed variables are summarized in Figure 3. Variables *Ttch hrs, Schoolenroll* and *Emppay* have the largest three missing rates, which are 76.7%, 66.9% and 60.5%, respectively.

**Figure 3:**
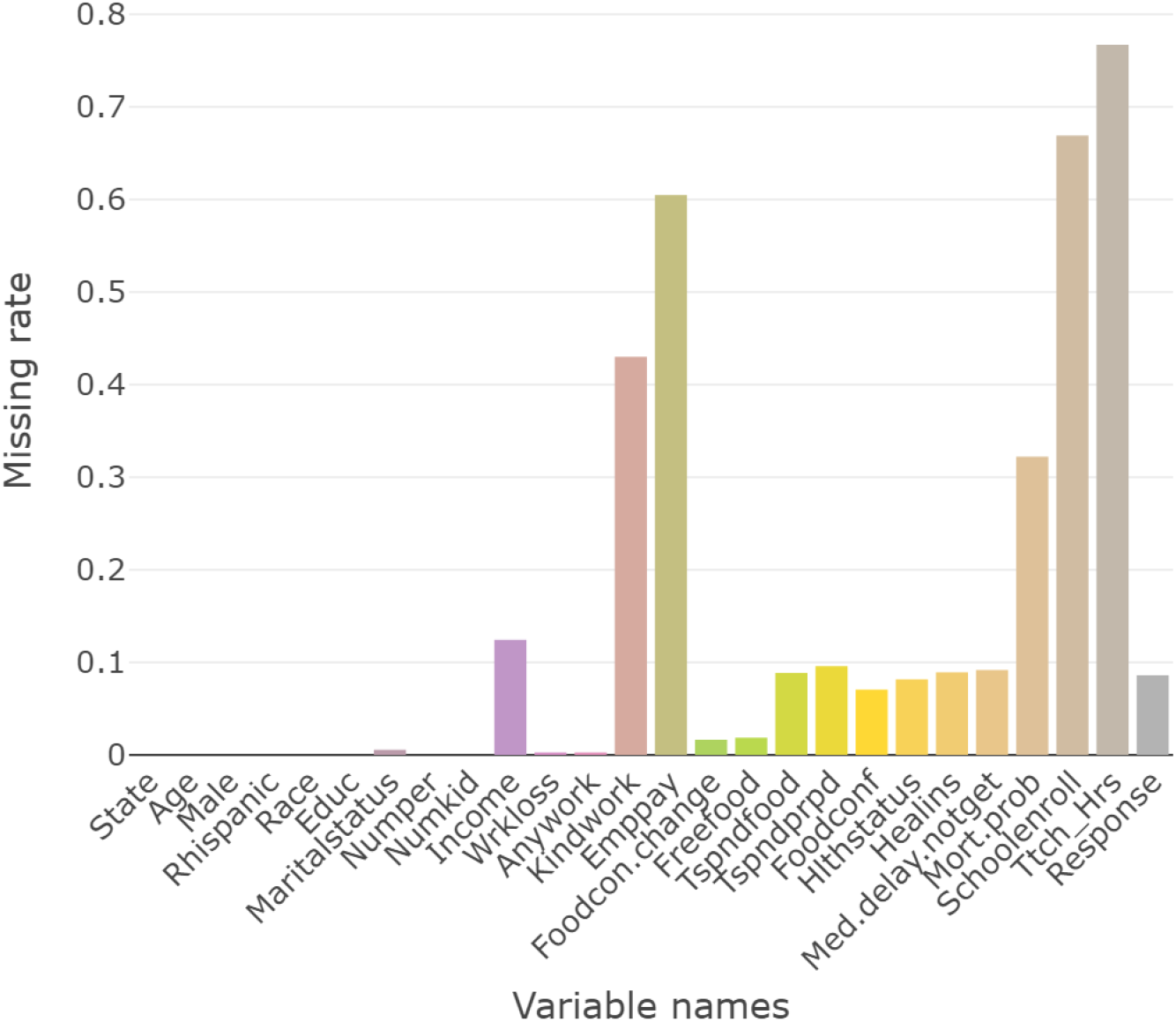
Missing rates for all the variables

### 3.1 Multiple Imputation by Chained Equations (MICE)

The presence of missing values in the dataset brings in a challenge for data analysis and model fitting. Leaving out the observations with missing features would not be the best strategy, and it would eliminate potential valuable information from the dataset or even yield biased results. A useful approach to handle missing observations in a complex dataset is *multiple imputation by chained equations* (MICE), which invokes fully conditional specification (FCS) under the assumption of the missing at random (MAR) mechanism. Each incomplete variable is imputed by its own imputation model which generates plausible values to replace the missing ones. MICE can be used for various types of variables with missing values, such as binary, continuous, nominal, and ordinal data. Technical details can be found in van Buuren et al. (2015).

### 3.2 Pre-Processing the Data with Imputation

Here we employ the MICE method to accommodate missing observations in the datasets. In each week, 5 distinct complete imputed datasets are created based on the original survey data by employing the same algorithm with different random seeds.

To compare the 5 imputed datasets to the original data for each week, we take week 6 as an example and show the distributions of the imputed data for both categorical and continuous variables. The density plots for three continuous variables *T spndf ood, Tspndprpd* and *Ttch hrs* are shown in Figure 4. The displays show that the distributions of the continuous variables in the imputed data are very similar to those in the original data. Three categorical variables *Anywork, Kindwork* and *Income* are selected as examples and their distributions are shown in Tables 3-5. Again, the proportions of different levels of each variable in five imputed datasets are fairly close to those in the original data.

**Table 3:** Proportions of different levels for *Anywork* in week 6

| Anywork | Level 1 | Level 2 |
| --- | --- | --- |
| Original Data | 0.5776 | 0.4224 |
| Imputation 1 | 0.5774 | 0.4226 |
| Imputation 2 | 0.5774 | 0.4226 |
| Imputation 3 | 0.5774 | 0.4226 |
| Imputation 4 | 0.5773 | 0.4227 |
| Imputation 5 | 0.5774 | 0.4226 |

**Figure 4:**
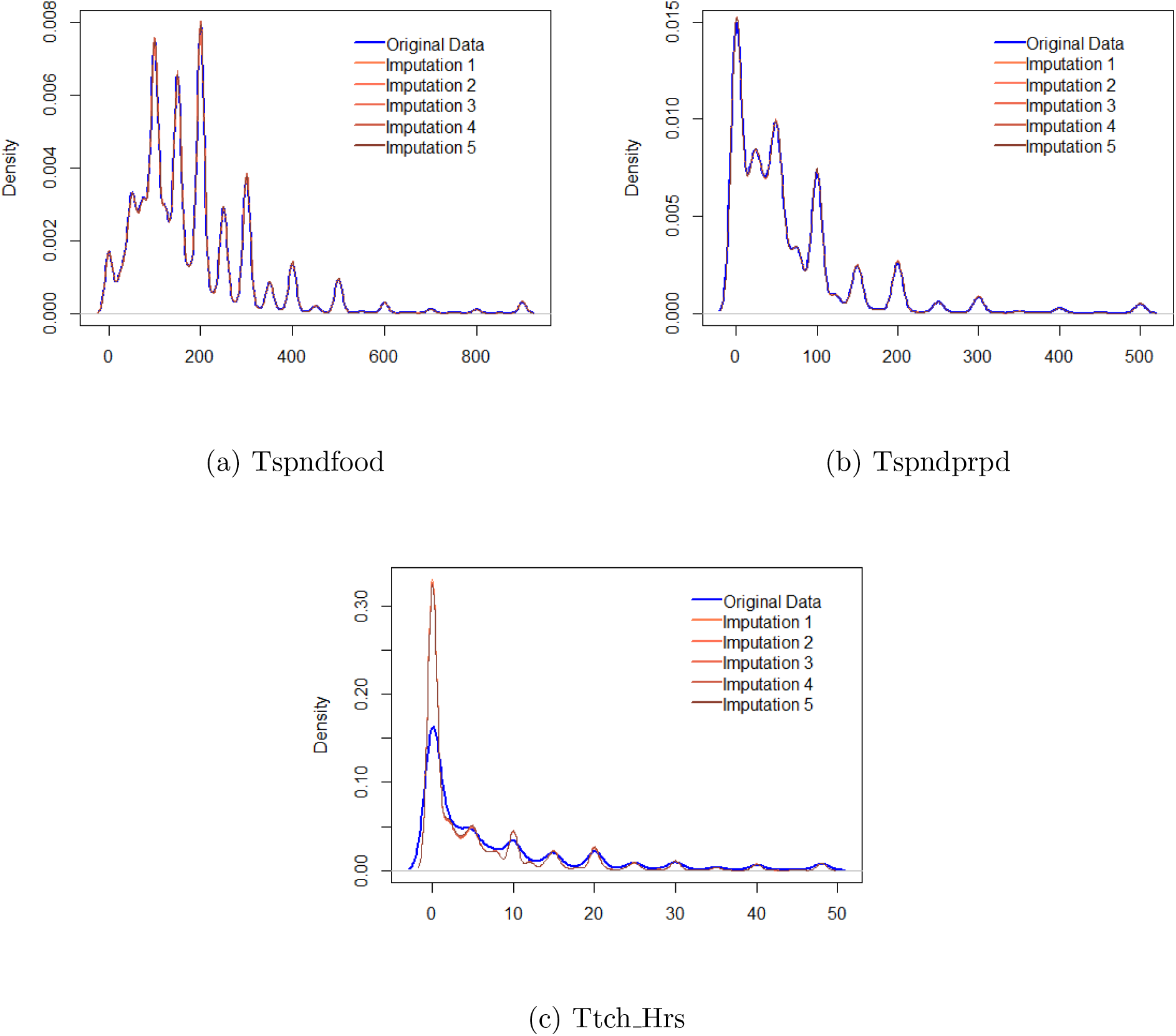
Density curves of the observed data and imputed data

**Table 4:**
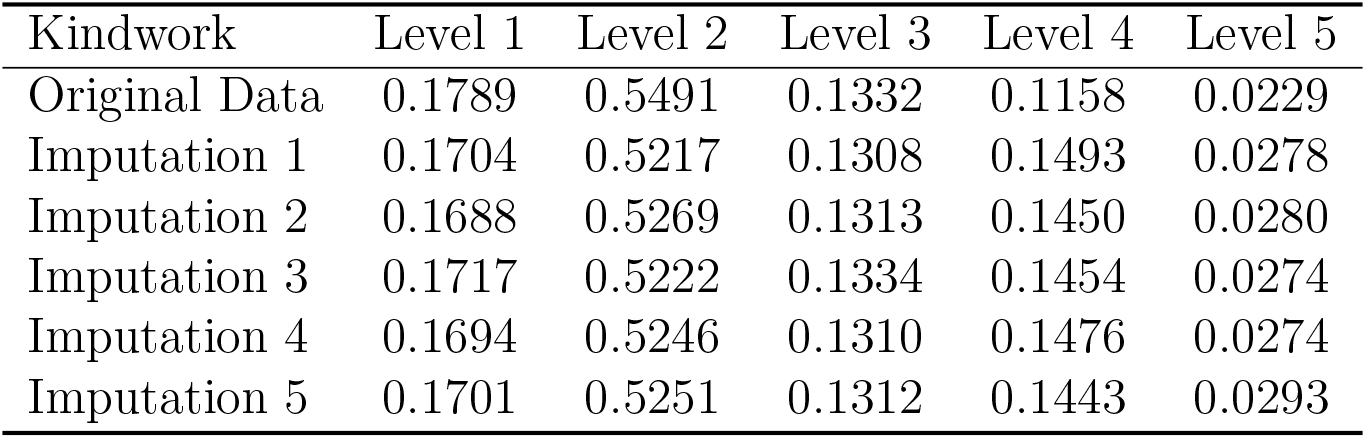
Proportions of different levels for *Kindwork* in week 6

**Table 5:** Proportions of different levels for *Income* in week 6

| Income | Level 1 | Level 2 | Level 3 | Level 4 | Level 5 | Level 6 | Level 7 | Level 8 |
| --- | --- | --- | --- | --- | --- | --- | --- | --- |
| Original Data | 0.1044 | 0.0876 | 0.1090 | 0.1765 | 0.1483 | 0.1858 | 0.0897 | 0.0986 |
| Imputation 1 | 0.1073 | 0.0892 | 0.1104 | 0.1779 | 0.1485 | 0.1829 | 0.0881 | 0.0958 |
| Imputation 2 | 0.1076 | 0.0892 | 0.1105 | 0.1782 | 0.1475 | 0.1833 | 0.0876 | 0.0961 |
| Imputation 3 | 0.1073 | 0.0896 | 0.1098 | 0.1773 | 0.1479 | 0.1844 | 0.0880 | 0.0957 |
| Imputation 4 | 0.1075 | 0.0897 | 0.1101 | 0.1773 | 0.1470 | 0.1842 | 0.0880 | 0.0962 |
| Imputation 5 | 0.1078 | 0.0892 | 0.1094 | 0.1772 | 0.1479 | 0.1843 | 0.0878 | 0.0964 |

## 4 Model Building and Inference

In this section, we employ the Lasso method to logistic regression to analyze the mental health data which contain a binary response and 25 predictors. For *i* = 1, …, *n*, let *Y*_*i*_ represent the binary response with value 1 indicating that the mental health problem occurs for subject *i* and 0 otherwise. Let *X*_*ij*_ denote the *j*th covariate for subject *i*, where *j* = 1, …, *p*, and *p* is the number of predictors. Write *X*_*i*_ = (*X*_*i*1_, *X*_*i*2_, …, *X*_*ip*_)^T^ and let *π*_*i*_ = *P* (*Y*_*i*_ = 1 |*X*_*i*_).

Consider the logistic regression model

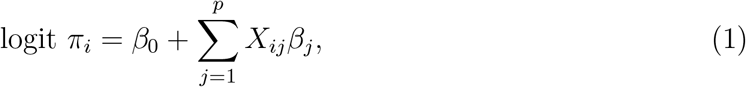

where *β* = (*β*_0_, *β*_1_, …, *β*_*p*_)^T^ denotes the vector of regression parameters.

The odds of the occurrence of mental health problems is defined by the ratio of the probability of having mental health problem happening to that of not having mental health issues i.e.,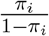 The log-likelihood function for *β* is given by

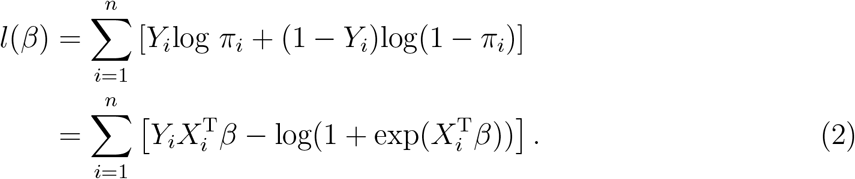

Since our objective is to select a subset of the predictors highly related to the dichotomous response, the Lasso method is used to do variable selection. The Lasso estimates are the values that maximize the penalized log-likelihood function, obtained by adding an *L*_1_ penalty to the function (2):

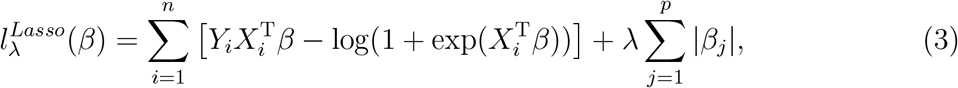

where *λ* is the tuning parameter that controls the complexity of the model; variable selection is realized by tuning the value of *λ*.

A proper value of the tuning parameter *λ* is data-driven and can be chosen by *K*-fold cross-validation, with *K* being user specified. In our analysis below, *K* is chosen as 10. We use the “*one-standard-error*” rule (Hastie et al., 2009, p. 60) to pick the most parsimonious model within one standard error of the minimum cross-validation misclassification rate. This rule was also used by other authors, such as Krstajic et al. (2014).

## 5 Analysis on the Mental Health Data

### 5.1 Model Fitting and Variable Selection

Each of the five imputed datasets obtained in Section 3.2 is considered as a complete surrogate of the original incomplete data. Now we apply the Lasso logistic regression to each imputed data for each week. The predictors corresponding to the nonzero coefficient estimates are the factors selected, which are basically different across five surrogate datasets for each of the 12 weeks. To have explorations in a full spectrum, we start with two extreme models, called the *full model* by including the union of all those selected factors, and the *reduced model* by including only the common factors selected for all five surrogate datasets in any week. The *full model* includes all the 25 predictors in the original data, and the *reduced model* contains 11 predictors: *Age, Male, MS, Numkid, Wrkloss, Anywork, Foodcon f, Hlthstatus, Healins, Med*.*delay*.*notget*, and *Mort*.*prob*. We expect the predictors in the *final model* to form a set in-between the sets of the predictors for the *reduced model* and the *full model*. Now, the problem is how to find the *final model* using the *reduced* and *full models*. To tackle this, we carry out the following steps.

In Step 1, we fit logistic regression with predictors in the *full model* and in the *reduced model*, respectively, to each of the five surrogate datasets for each of the 12 weeks. In Step 2, the estimates and standard errors of the model coefficients for a given week are obtained using the algorithm described by Allison (2000). To be specific, let *M* be the number of surrogate datasets for the original incomplete data, which is 5 in our analysis. Let *β*_*j*_ be the *j*th component of the model parameter vector *β*. For *k* = 1, …, *M*, let 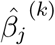 denote the estimate of the model parameter *β*_*j*_ obtained from fitting the *k*th surrogate dataset in a week and let 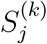 be its associated standard error. Define

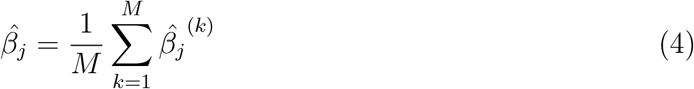

and

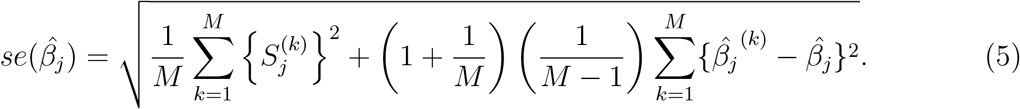

Then 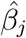 is taken as the point estimate of *β*_*j*_, and 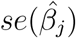 is used as the associated standard error. The results of the *full* and *reduced models* for the 12 weeks are displayed in Tables 6 and 7, respectively.

**Table 6:**
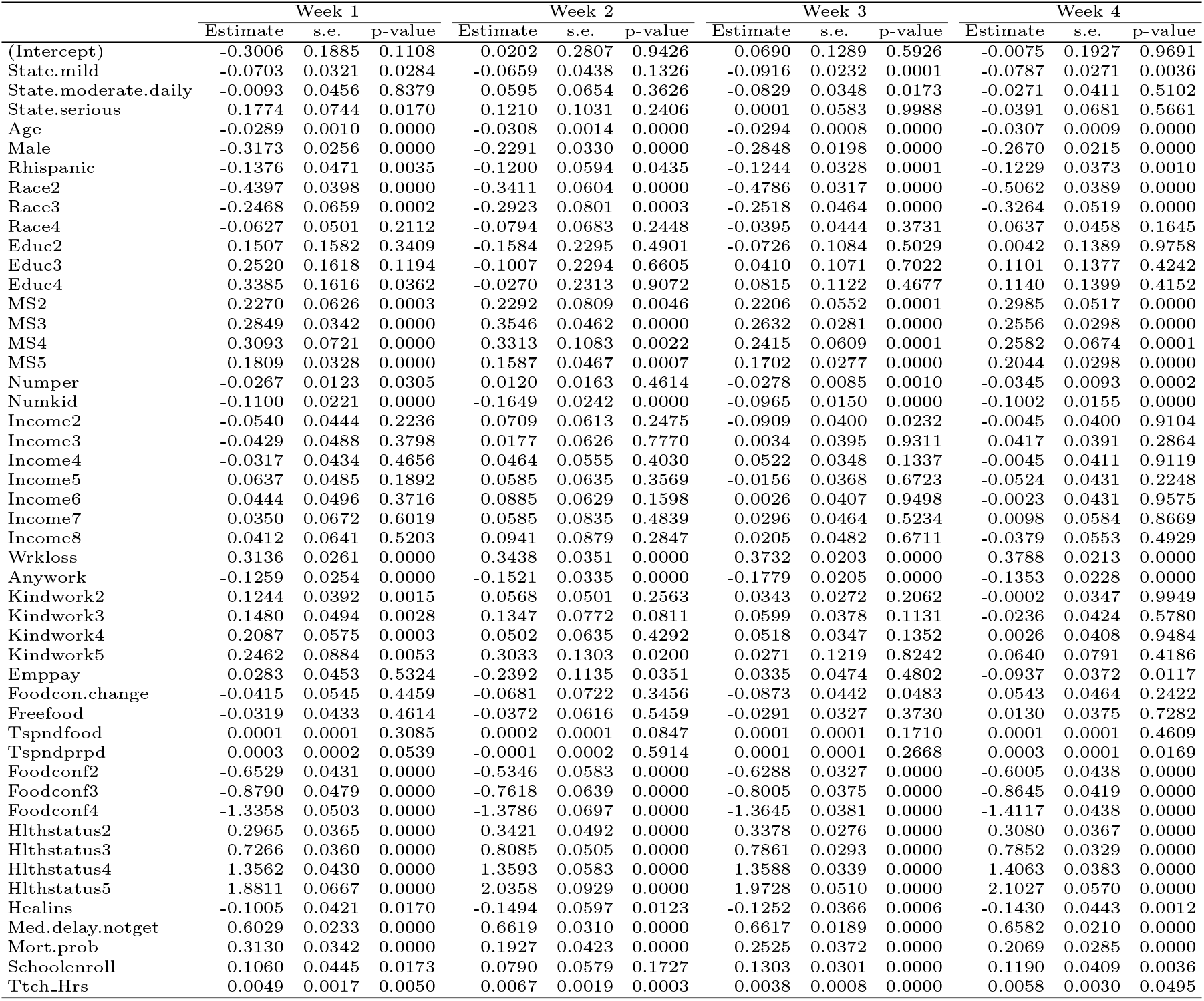
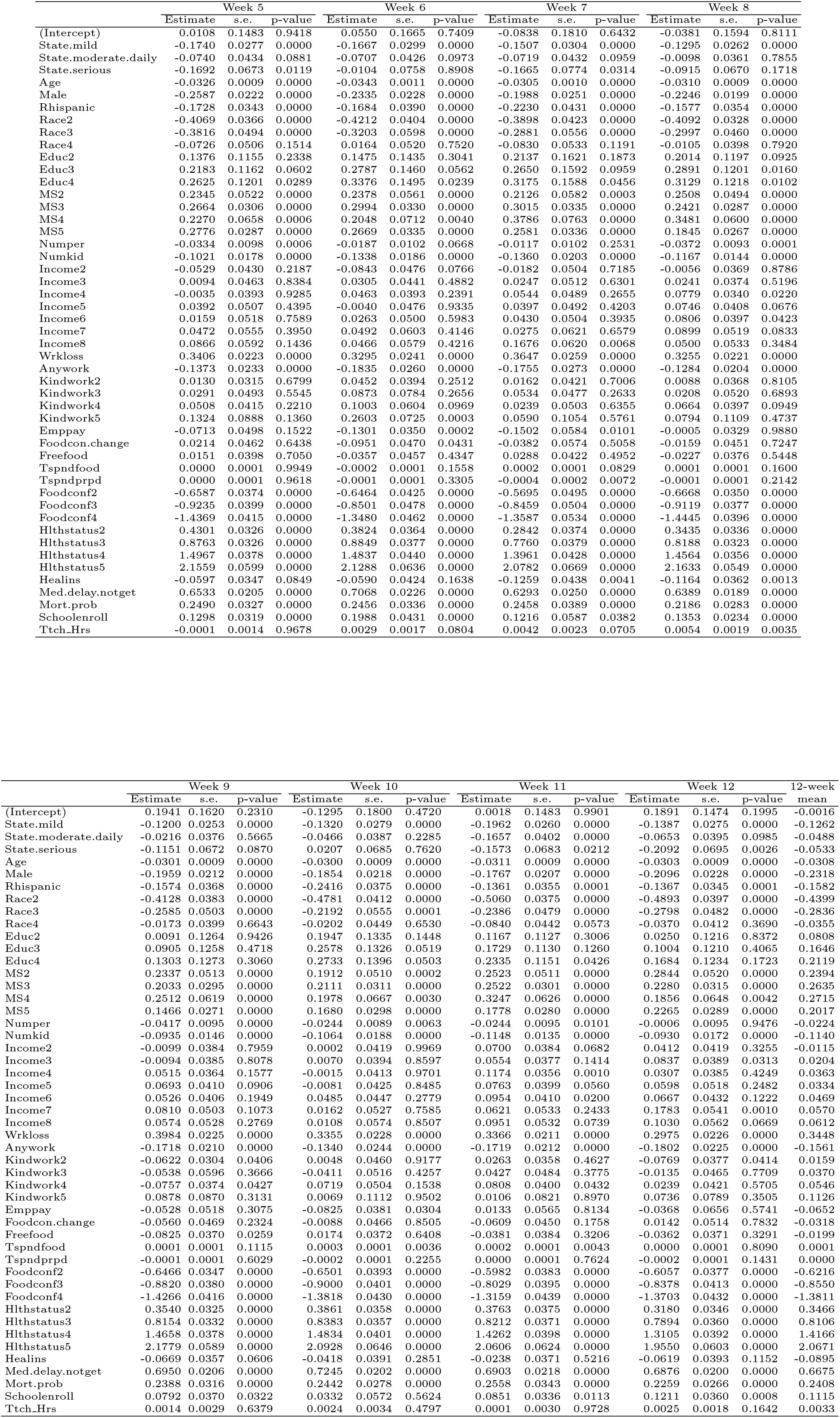
Coefficient estimates, standard errors and p-values for the *full model*

**Table 7:**
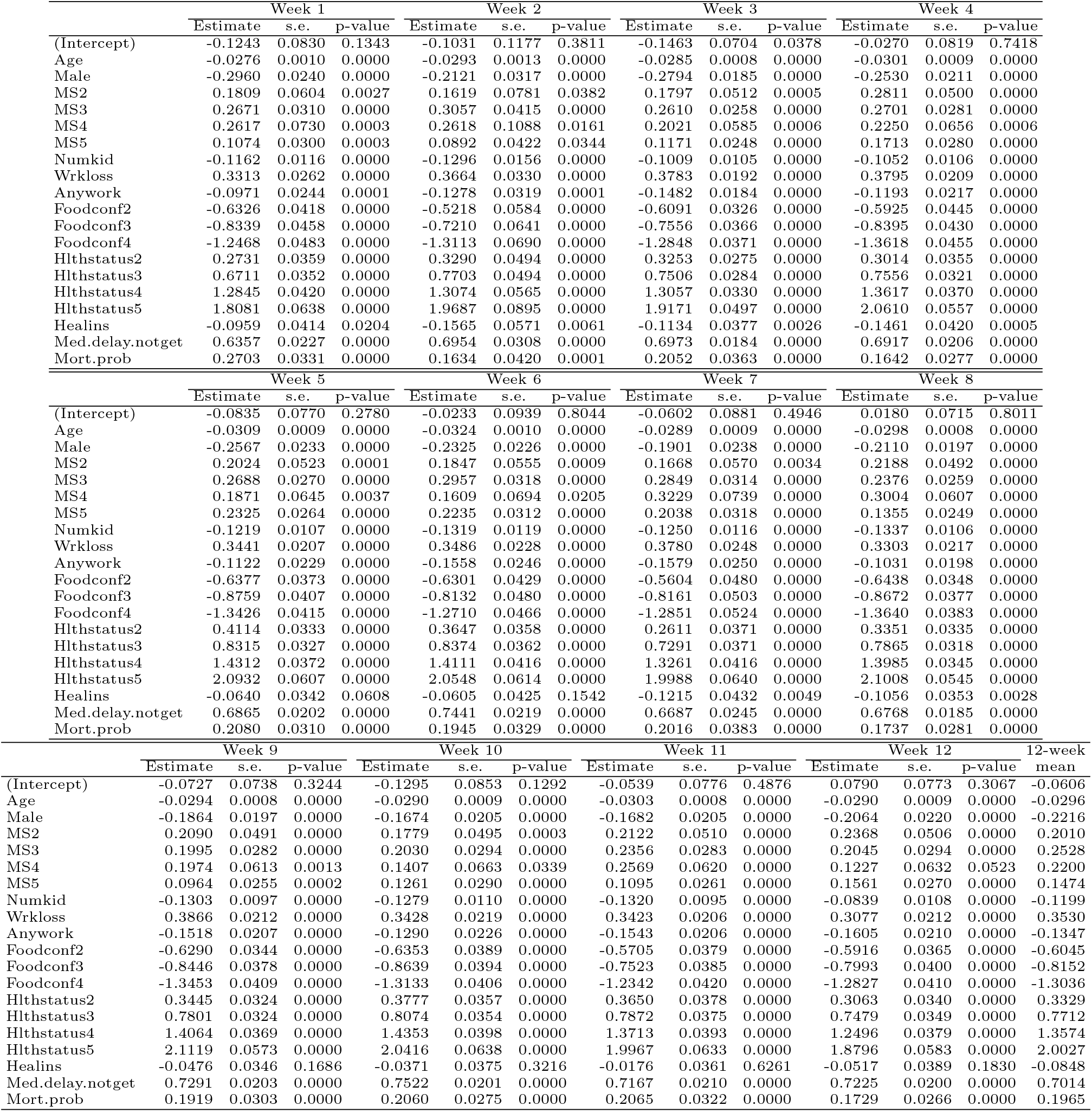
Coefficient estimates, standard errors and p-values for the *reduced model*

In Step 3, we combine those 11 predictors in the *reduced model* with the five additional variables in the *full model* that are significant (under the significant level 0.05) in the analysis from Step 2 for at least 6 weeks’ data, which are *State, Rhispanic, Race, Numper*, and *Schoolenroll*.

In Step 4, we construct the *final model* by using the model form (1) to include the selected variables in Step 3 as predictors. To do so, we re-express discrete variables with multiple categories using dummy variables. *State, Race, MS, Foodconf*, and *Hlthstatus* are expressed using 3, 3, 4, 3, 4 dummy variables, respectively. That is, the *final model* is given by

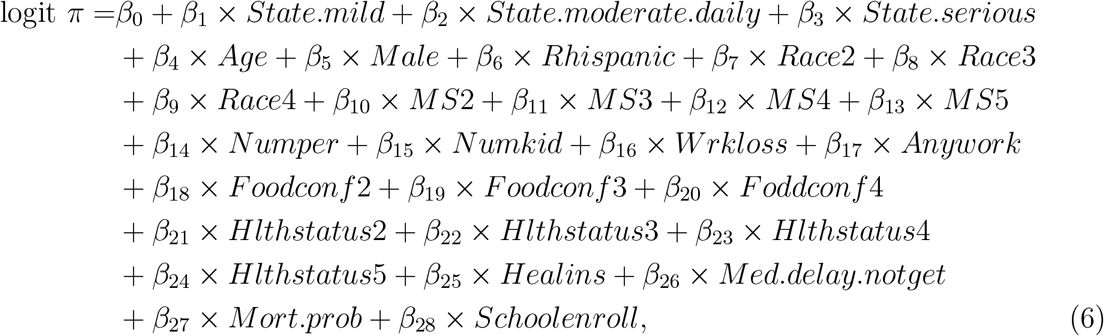

where *β*_*j*_ is the regression coefficients for *j* = 0, 1, …, 28.

Then, we fit the final logistic model (6) to the surrogate datasets for each of the 12 weeks. Using the algorithm of Allison (2000) again, we obtain the point estimates of the model parameters and the associated standard errors in the same manner as indicated by

(4) and (5). The results are reported in Table 8.

**Table 8:**
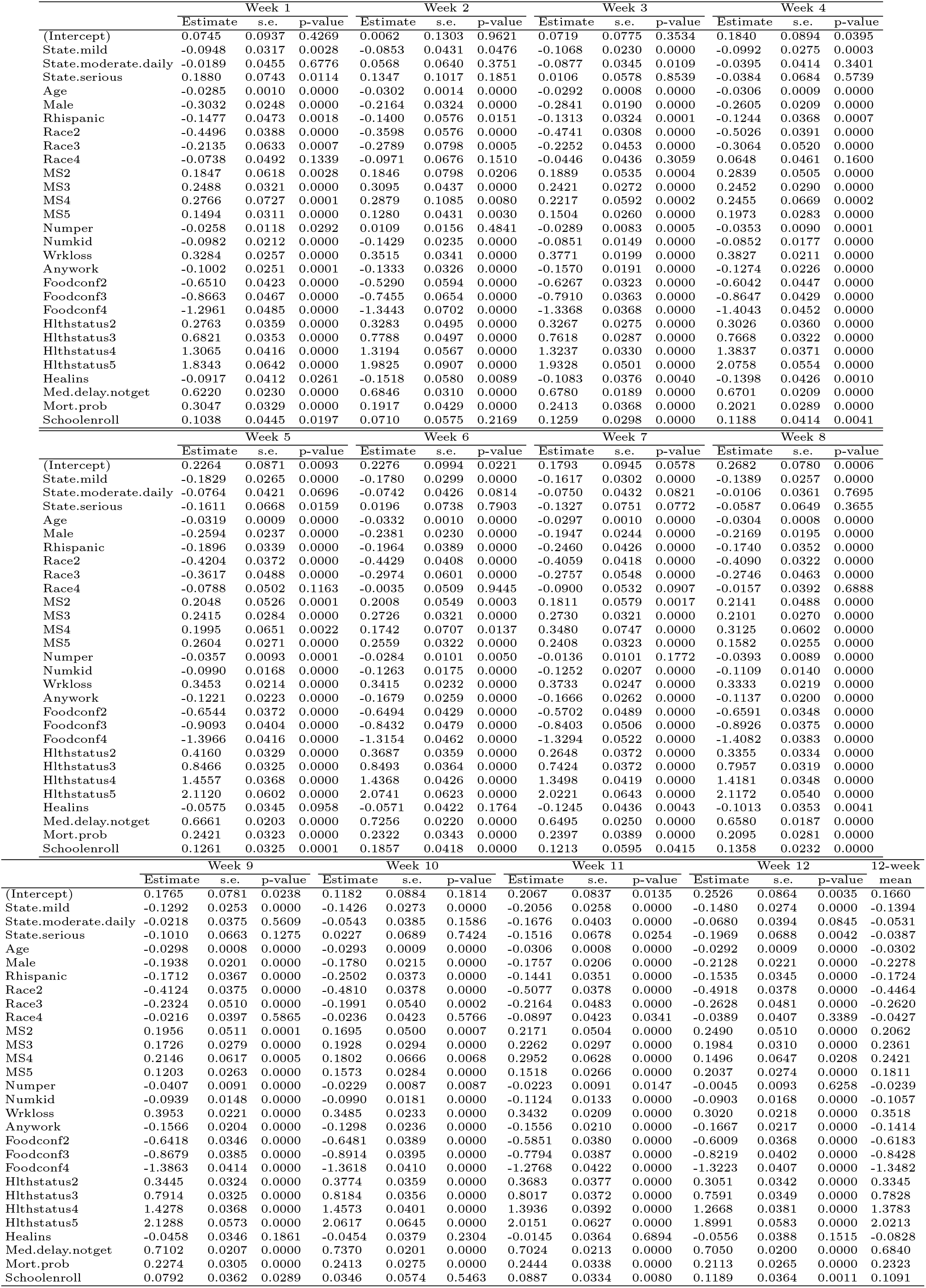
Coefficient estimates, standard errors and p-values from the *final model* for the 12 weeks

### 5.2 Numerical Findings

For intuitive visualization, we plot the estimates for the significant coefficients for the 12 weeks in Figure 5. It is seen that the absolute value of coefficient estimates for some levels of variables *Hlthstatus* and *Foodconf* are greater than 1. The coefficient estimates of *Med*.*delay*.*notget* along 12 weeks are between 0.5 and 1. Other variables have coefficient estimates between −0.5 and 0.5.

**Figure 5:**
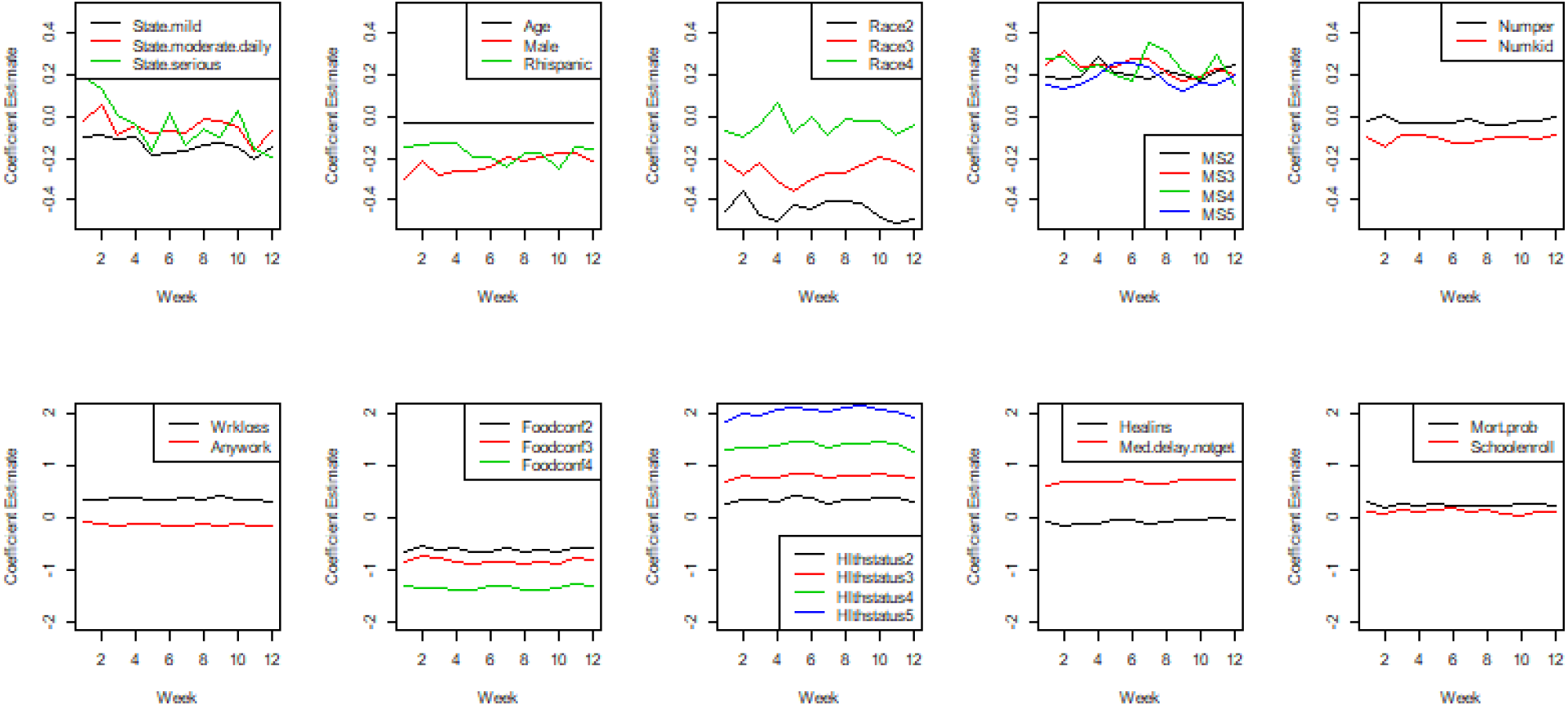
Coefficient estimates for the 12 weeks

Next, we interpret the average effects of the predictors for the 12 weeks. We first look at the estimate results for *State*. Considering the States with large daily increases of cases as the baseline and on the average of 12 weeks, people from mild pandemic States exhibit a 13.01% decrease in the odds of having mental issues; people from the States with small daily increases show a 5.17% decrease in the odds; people from serious pandemic States are generally associated with a 3.80% decrease in the odds. It is worthwhile to point out that the coefficient estimates for *State*.*mild* have a decreasing trend along 12 weeks (shown in Figure 5), with the absolute values increase. This indicates that the pandemic effect on the mental health problems in the States with mild pandemic increase as time goes by.

For *Age* and *Gender*, the results indicate that the increase in age and being a male relative to a female are generally associated with a decrease in the odds of having mental issues. Specifically, the 12-week mean of the coefficient estimates of *Age* and *Male* are −0.0302 and −0.2278, respectively. It means that, on average, one unit increase in *Age* is associated with about a 1 *−* exp(*−*0.0302) *≈* 2.97% decrease in the odds of occurrence of mental health problems; and being a male relative to a female is associated with a 1 *−* exp(—0.2278) ≈ 20.37% decrease in the odds of having mental health issues. Interestingly, there is an increasing trend of the coefficient estimate of males along 12 weeks (as shown in Figure 5), suggesting that the gender effect on the occurrence of mental health problem decreases as time goes by. Similarly, the overall 12-week effects of *Rhispanic* indicate that the origin of Hispanic, Latino or Spanish is associated with a smaller odds of having mental issues than others. The 12-week mean of the coefficient estimates of *Rhispanic* is −0.1724, leading to the odds of mental health problem occurrence being reduced by around 1 − exp(— 0.1724) ≈ 15.84%.

For variable *Race*, considering the White people as the baseline, the Black and the Asian tend to have less odds of getting mental health problems. The 12-week mean of coefficient estimates for the Black and the Asian are −0.4464 and −0.2620, respectively, yielding that the odds of occurrence of mental health issues for the Black and the Asian are 1 — exp(— 0.4464) ≈ 36% and 1 − exp(− 0.2620) ≈ 23.05% less relative to the White people.

For *MS* (Marital status), considering *now married* as the baseline, the results suggest that people who are *widowed, divorced, separated*, or *never married* are associated with a 22.90%, 26.63%, 27.39%, and 19.85% increase in the odds of having mental issues, respectively, relative to the people who are *now married*.

For predictors *Numper* and *Numkid*, the results show that the increase of the number of people and kids in the household is associated with the decrease of the odds of having mental issues. Specifically, one person increase in the household is associated with 2.36% decrease in odds, and one more kid in the household is associated with 10.03% decrease in the odds.

For the work-related factors *Wrkloss* and *Anywork*, the results indicate that experiencing a loss of employment income since March 13, 2020 is associated with a 42.16% increase in the odds of having mental issues, and doing any work during the last 7 days is associated with a 13.19% decrease in the odds.

The results of *Foodconf* show that an increase in the confidence of food affordability is negatively associated with the odds of having mental issues. On average of 12 weeks, the more confident in the food affordability, the less the odds of having mental issues. For example, people confident in the food affordability for the next four weeks demonstrate a 74.03% decrease in the odds of having mental issues.

For *Hlthstatus*, using the health condition *excellent* as the baseline, the results say that the worse the self-evaluated health condition, the larger the odds of having mental issues. Considering the worst level of health condition *poor* as an example, people who think they are in *poor* health conditions have an odds of having mental issues 7.55 times higher than that of *excellent* people.

The results for other health-related predictors, *Healins* and *Med*.*delay*.*notget*, indicate that, on average of 12 weeks, people who are currently covered by health insurance are associated with a 7.95% decrease in the odds of mental issues occurrence, and people who are not get medical care or have delayed medical care are generally associated with a 98.18% increase in the odds.

The estimates on *Mort*.*prob* and *Schoolenroll*, on average of 12 weeks show that people having rental or mortgage problems are associated with a 26.15% increase in the odds of having mental health problems, and that people whose household has kids enrolled in school are associated with a 11.53% increase in the odds of having mental issues.

In summary, the factors associated with a reduction in the odds of having mental health problems are: States not having large daily increases of cases, the increase of age, being male, having a Hispanic, Latino or Spanish origin, being non-White, the increase in number of people or kids in the household, having job during the last 7 days, having confidence in the food affordability in the future, and being covered by insurance. The factors associated with the increase in the odds of getting mental issues are: not married, experiencing loss of job, poor self-evaluations on the health condition, having problems in getting medical care and mortgage, and having kids enrolled in school. Among all the predictors in the *final model, State* and *Male* are two predictors having apparent trends along 12 weeks. Specifically, the effect of *State* decreases and the effect of gender increases over time.

## 6 Discussion

This paper quantitatively investigates the impact of the COVID-19 pandemic on the public mental health issues using the data in the United States for the period of April 23, 2020 −July 21, 2020. Multiple imputation and data pre-processing procedures are implemented to account for the effects due to missingness and the error-prone values in the original data. We employ the penalized logistic regression with the Lasso penalty to identify significant risk factors on mental health issues. Our analysis shows that health-related factors and confidence in the future food are important predictors related to the occurrence of mental health problems. The effects of most of the predictors are fairly stable along 12 weeks, except for States and age.

While this study offers us quantitative evidence how the COVID-19 pandemic can psychologically challenge the public, several limitations need our future explorations. Firstly, the data used in this project are from an online survey studying how the coronavirus pandemic is impacting households across the US from social and economic perspectives. Thus, the data may not contain enough necessary factors related to mental health issues. Secondly, the interaction effects between the predictors are not considered in our analysis, so the capacity of the model may be somewhat limited. Lastly, while an imputation method is used to handle missing observations, due to the large proportion of missing values in the data, the analysis results incur extra variability induced from the additional data management procedure.

The analysis here provides us evidence-based findings on the pandemic impact on the public mental health, which, in turn, offers us the guidance on the prevention of people from having negative emotions, such as depression, anxiety, and stress. Basically, it is very important to improve the welfare and take actions to ensure the public health care, for example, trying to improve the proportion of people covered by the insurance. Secondly, improving public confidence in future food conditions is also crucial. Lastly, actions and policies should be adopted to deal with the current low employment rate. In the end, more psychological counseling and psychotherapy services should be provided to the public to help them face the great change in the life because of the COVID-19 pandemic.

## Data Availability

The data used are publicly accessible.

https://www.census.gov/

## Acknowledgements

The research was supported by grants from the Natural Sciences and Engineering Research Council of Canada. Yi is Canada Research Chair in Data Science (Tier 1). Her research was undertaken, in part, thanks to funding from the Canada Research Chairs program.

## Notes

### Competing Interest Statement

The authors have declared no competing interest.

